# Avoidable burden of stomach cancer and potential gains in healthy life years from gradual reductions in salt consumption in Vietnam, 2019 – 2030: a modelling study

**DOI:** 10.1101/2022.02.12.22270881

**Authors:** Leopold Ndemnge Aminde, Linda J Cobiac, Dung Phung, Hai N Phung, J. Lennert Veerman

## Abstract

**Objective:** Excess salt consumption is causally linked with stomach cancer. However, the impact of high salt intake on stomach cancer in Vietnam is not known. The aim of this study was to quantify the future burden of stomach cancer that could be avoided from population-wide salt reduction in Vietnam.

**Design:** A dynamic simulation model was developed to quantify the impacts of achieving the 2018 National Healthy Vietnam program (8g/day by 2025 and 7g/day by 2030) and the WHO (5g/day) salt reduction targets. Data on salt consumption were obtained from the Vietnam 2015 WHO STEPS survey. Health outcomes were estimated over 6-year (by 2025), 11-year (by 2030) and lifetime horizons. We conducted one-way and probabilistic sensitivity analyses.

**Setting:** Vietnam

**Participants:** All adults aged ≥ 25 years (total of 61 million people, 48.4% men) alive in 2019.

**Results:** Achieving the 2025 and 2030 national salt targets could result in 3,500 and 7,700 fewer incident cases of stomach cancer respectively, and avert 1,950 and 5,200 stomach cancer deaths, respectively. Achieving the WHO target by 2030 could prevent 9,280 new cases of, and 6,300 deaths from stomach cancer. Over the lifespan, this translated to 359,000 (8g/day), 416,000 (7g/day) and 505,000 (5g/day) HALYs gained respectively.

**Conclusions:** A sizeable burden of stomach cancer could be avoided, with gains in healthy life years if national and WHO salt targets were attained. Our findings provide impetus for policy makers in Vietnam and Asia to intensify salt reduction strategies to combat stomach cancer and mitigate pressure on the health systems.

## Introduction

Stomach cancer – also known as gastric cancer – is a major public health problem in many countries. In 2019, there were over 1.2 million newly diagnosed cases of stomach cancer worldwide. It is the fifth most commonly diagnosed cancer globally and the third leading cause of cancer mortality (1). In 2019, stomach cancer was responsible for almost a million deaths and accounted for over 22 million disability adjusted-life years (DALYs) deaths (2). The largest share of this cancer burden occurred in low-income and middle-income countries (LMICs), where most healthcare systems are ill-equipped to provide the complex care that is required.

Vietnam is among the top ten countries with the highest rates of stomach cancer worldwide (1, 2). Stomach cancer is the third most frequently diagnosed cancer in Vietnam, and in 2018, over 17,500 new cases of stomach cancer were diagnosed in the country. It is also the third leading cause of cancer mortality in Vietnam (3). The five-year survival rate of people with stomach cancer is very poor, typically ranging between 20 and 30%. (2, 4). With such grim prognosis, primary prevention is crucial, especially in LMICs.

High salt (sodium) consumption is thought to increase the risk for stomach cancer via multiple mechanisms. Studies in animals have demonstrated that chronic exposure to high salt intake alters the viscosity of the mucous lining of the stomach, progressively causing atrophic gastritis and intestinal metaplasia. In addition, high salt concentrations facilitate *Helicobacter pylori* colonization of the stomach, which then upregulates the expression of the carcinogenic protein (cagA) (5). Exposure to high salt diets also promotes the formation of N-nitroso compounds, which are well-known human carcinogens associated with gastrointestinal cancers (6, 7). Pooled evidence from prospective cohort studies have demonstrated that high salt consumption significantly increases the risk of developing stomach cancer (8, 9). A recent dose-response meta-analysis by Fang and colleagues found that a 5g/day increment in dietary salt intake increased the risk of stomach cancer by 12% (9). Given very high average dietary salt intake globally, this translated to attribution of well over a third (38.2%) of the stomach cancer DALYs to diets high in sodium in the recent Global Burden of Disease (GBD) analysis (2).

The most recent World Health Organization (WHO) STEPS survey in Vietnam estimated that adults consume on average 9.4 grams of salt per day (10), which is about double the current WHO recommendation of <5g/day (11). Existing research on the harmful effects of high salt intake globally (12) and in Vietnam (13, 14) has mostly focused on its impact on blood pressure and cardiovascular disease. The impact of such high salt diets on the burden of stomach cancer has not been evaluated in Vietnam. Recognizing high salt consumption as a major public health concern, the government of Vietnam in their recent ‘Healthy Vietnam Program’ has set national average salt reduction targets to be achieved by 2025 and by 2030 (15). Building on the above evidence gaps, the aim of this study was to: 1) Quantify the avoidable incidence of and mortality from stomach cancer and; 2) Estimate the impact of these changes in stomach cancer burden on healthy life years, if the Healthy Vietnam Program and WHO salt reduction targets were achieved.

## Methods

### Study design, population, and data sources

We conducted a simulation study using a multi-cohort proportional multistate lifetable (PMSLT) model. This model combines demographic, epidemiological and economic data to project the impacts of interventions over the lifecourse and is well-suited for population-level chronic disease modelling given its capacity to account for comorbidities (16, 17). All adult Vietnamese aged ≥ 25 years (total of 61 million people, 48.4% men) alive in 2019 were included in this analysis.

Age- and gender-specific data on salt intake was obtained from the 2015 Vietnam WHO STEPwise approach to NCD surveillance (STEPS) study (10). This nationally representative survey included adults aged 18 to 69 years from 4,651 households across all 63 provinces in Vietnam. Spot urine samples were collected from participants to measure urinary sodium (salt) content. The INTERSALT formulae (18) were then used to estimate 24-hour urinary sodium excretion from the spot urine samples, and these were used as a proxy for sodium intake in this study. Due to the absence of reliable data, we assumed people ≥ 70 years had similar salt intake levels to those aged 69 years. For baseline stomach cancer epidemiology, age- and sex-specific data was obtained from the 2019 GBD study. Model input parameters are provided in the appendix.

### Modelling framework

The PMSLT is a dynamic model that simulates age- and sex-stratified cohorts of the Vietnamese population ≥ 25 years alive in 2019. The model has two main populations – a reference population with current risk factor distribution, baseline stomach cancer epidemiology and population demographics, and an identical population that receives the ‘intervention’.

#### Modelling salt intake and stomach cancer risk

Dietary salt intake was modelled as a continuous distribution. The age- and sex-stratified population salt distributions were shifted from current levels towards achieving the mean intake levels stipulated in the Healthy Vietnam Program (8g/day by 2025 and 7g/day by 2030) and the WHO recommendation (5g/day) by 2030. Estimates of the direct link between salt intake and stomach cancer risk were obtained from the GBD 2019 comparative risk assessment study (19). For the main analysis, we considered the optimal salt distribution or theoretical minimum risk exposure level (TMREL) to be 3 grams of sodium (∼7.6g of salt) and varied this to 1g and 5g of sodium in the sensitivity analysis GBD (20). By integrating the exposure distribution and the risk function in the potential impact fraction (PIF) – which calculates the proportional change in a disease condition following a change in the exposure or risk factor, we estimate the post-intervention incidence of stomach cancer. See Box 1.

##### Box 1

**Potential impact fraction formula and post-intervention incidence calculation**

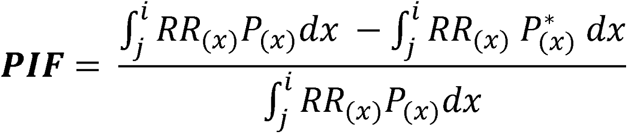

Where, x = Salt intake levels, RR_(x)_ = relative risk, P_(x)_ = baseline salt distribution, P* = salt distribution after the intervention, i = start integration level (0.5g Salt), j = end integration level (25g Salt). dx = integration with respect to x, with i and j as the integration boundaries.

The formula: **I’ = I**_**\***_**(1-PIF)**, where I’ = incidence after intervention, I = baseline incidence; is used to calculate the post-intervention stomach cancer incidence.

#### Stomach cancer state-transition and lifetable modelling

The stomach cancer state-transition model contains four states: Healthy (alive and free from stomach cancer), Diseased (alive with stomach cancer), Dead from stomach cancer and Dead from other causes. Annual probabilities of proportions of the population transiting between these states is influenced by incidence, remission and case fatality hazards (*Figure 1*). The PIF described above feeds into the ‘intervention’ section of the stomach cancer state-transition model to estimate post-intervention incidence, which influences the prevalence and then mortality rates. The latter feed into the ‘intervention’ lifetable, altering the all-cause mortality rates, and propagate through to estimate life years. These life years (LY) are adjusted for poor quality of life to obtain health-adjusted life years (HALY). This quality-of-life adjustment is obtained by dividing the age- and sex-stratified years lived with disability (YLD) for stomach cancer by the prevalence of stomach cancer, and then adjusted for background disability using all-cause population YLD. The sex-stratified five-year age cohorts of the population are simulated through annual cycles until everyone reaches 100 years or dies. The impact of the intervention is determined by the difference in incidence, mortality and HALYs between the reference and the intervention populations.

**Figure 1:**
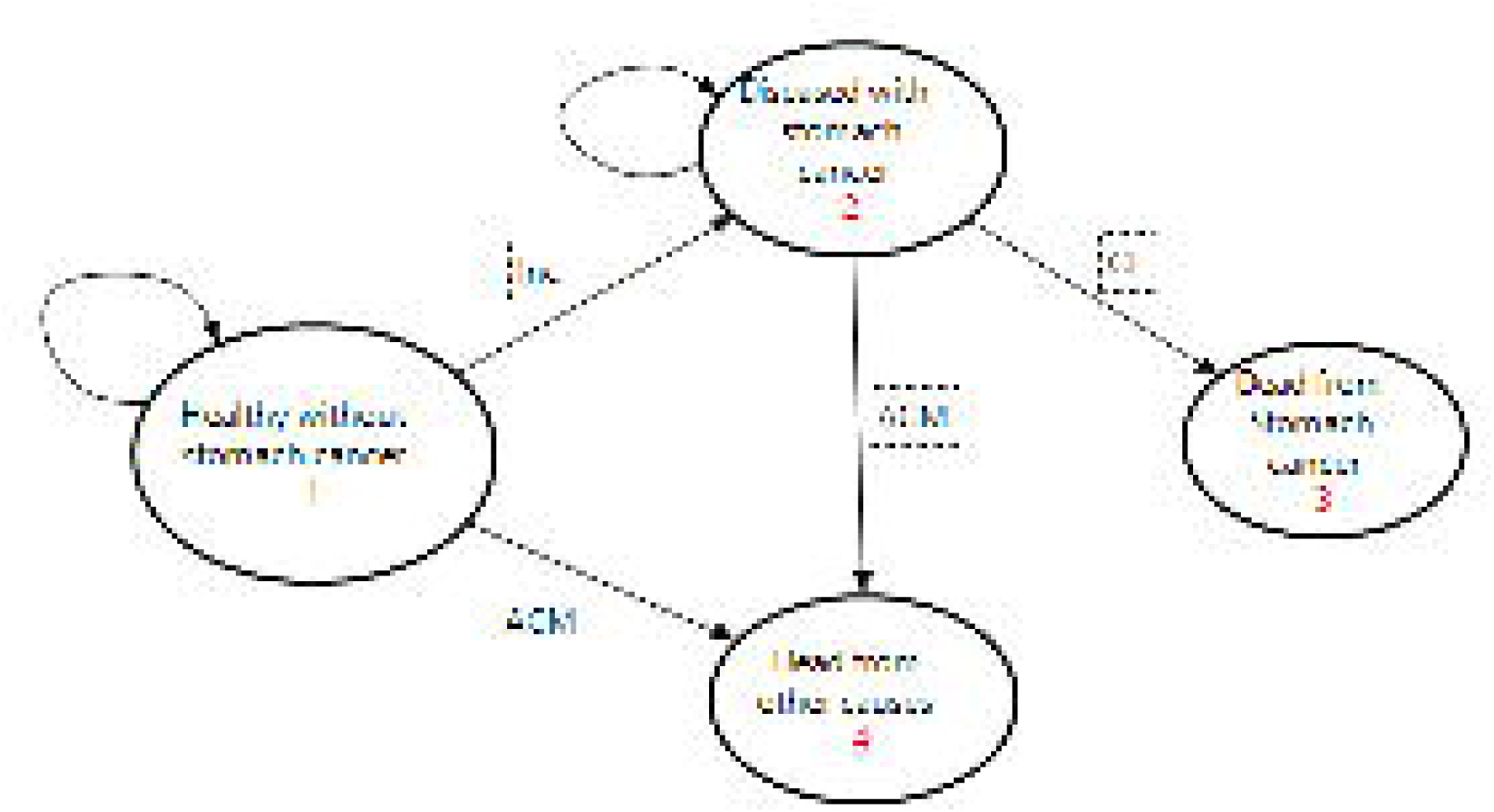
State-transition diagram showing the four health states for the stomach cancer Markov model. Inc = Probability of stomach cancer incidence, CF = case fatality probability, ACM = All-cause mortality probability. Straight arrows imply for each annual cycle, the progression (transition) of proportions of the population from each sex and age-group cohort to the next state, while the circular arrows imply probability of remaining in the same state.

### Scenarios and sensitivity analysis

In the base case analysis, we quantified the changes in health outcomes following a gradual (linear) reduction in salt consumption to *i)* an average of 8g/day by 2025 (National Target [NT] 1); *ii)* an average of 7g/day by 2030 (NT 2); and *iii)* an average of 5g/day as recommended by WHO by 2030 (WT). In further scenarios, we modelled lifetime impacts, that is, the national and WHO targets achieved and sustained for the remaining lifetime of the population. Next, we modelled intervention decay scenarios whereby: NT 1 levels are achieved and sustained for the first five years (until 2030) followed by a cumulative quarterly reduction in effect every five years until zero impact beyond the year 2045. In a like manner, NT 2 and WT are achieved and sustained for the first five years (until 2035) followed by a cumulative quarterly reduction in effect every five years until zero impact beyond the year 2050. Current evidence appears unsettled on what the optimal level or TMREL of sodium (salt) intake should be, that is, the level of consumption at which disease risk is lowest (20). In our base case, we used a TMREL of 3g of sodium (∼7.6g of salt) consistent with the GBD study and conducted one-way sensitivity analysis using 1g and 5g of sodium as optimal levels (19, 20).

### Uncertainty analyses

To assess parameter uncertainty in our model, we attributed plausible statistical distributions to key parameters: salt intake levels (normal distribution) and relative risks (lognormal distribution). Probabilistic sensitivity analyses using second-order Monte Carlo simulations were conducted to jointly capture the overall uncertainty of key parameters in our outcome projections. We ran 2,000 simulations and report the 95% uncertainty intervals (2.5th and 97.5th percentile) around our best estimates. The simulations were implemented using the Ersatz 1.35 software (Epigear International, Australia) (21).

## Results

### Estimated population salt reduction

We modelled a gradual reduction in average population salt intake from the 2019 base year level to the policy targets. For 25–30-year-old men, this was an estimated mean salt reduction of 0.383g per annum (to reach 8g/day by 2025), 0.300g per annum (to reach 7g/day by 2030) and 0.482g per annum (to reach 5g/day by 2030). Corresponding estimates for women were 0.033g, 0.109g and 0.291g per annum, respectively. Table 1 in the supplementary file depicts the baseline salt intake and the estimated average age- and sex-stratified annual reductions modelled.

**Table 1:** Estimated reductions in incidence of stomach cancer for adults in Vietnam between 2019 to 2030 and lifetime.

| New cases | Men |  | Women |  | Both |  |
| --- | --- | --- | --- | --- | --- | --- |
|  | *Absolute reduction*<br>*n (95% UI)* | *Relative reduction*<br>*Est. (95% UI)* | *Absolute reduction*<br>*n (95% UI)* | *Relative reduction*<br>*Est. (95% UI)* | *Absolute reduction*<br>*n (95% UI)* | *Relative reduction*<br>*Est. (95% UI)* |
| NT 1a | 2,902 | 6.2 | 597 | 2.8 | 3,499 | 5.1 |
|  | (1,109 - 5,199) | (2.4 - 11.1) | (133 – 1,316) | (0.6 - 6.1) | (1,589 – 5,840) | (2.3 - 8.5) |
| NT 2a | 6,354 | 7.3 | 1,346 | 3.3 | 7,701 | 6.0 |
|  | (2,790 - 11,030) | (3.2 - 12.6) | (354 – 2,688) | (0.9 - 6.7) | (3,880 – 12,684) | (3.0 - 9.9) |
| WT a | 7,546 | 8.6 | 1,732 | 4.3 | 9,278 | 7.3 |
|  | (3,969-11,916) | (4.5 - 13.6) | (682 – 3,133) | (1.7 -7.8) | (5,431 – 13,959) | (4.3 - 10.9) |
| **Lifetime – with decay effects** |  |  |  |  |  |  |
| NT 1b | 15,392 | 3.3 | 2,910 | 1.1 | 18,302 | 2.5 |
|  | (6,963 – 26,329) | (1.5 - 5.7) | (668 – 6,404) | (0.3 - 2.5) | (9,103 – 28,999) | (1.3 - 4.0) |
| NT 2b | 21,782 | 4.7 | 4,529 | 1.8 | 26,310 | 3.6 |
|  | (10,264 – 35,773) | (2.2 - 7.7) | (1,340 – 8,975) | (0.5 - 3.5) | (13,892 – 41,175) | (1.9 - 5.7) |
| WT b | 26,376 | 5.7 | 5,670 | 2.2 | 32,046 | 4.4 |
|  | (15,101 – 40,014) | (3.3 - 8.6) | (1,913 – 10,238) | (0.7 - 4.0) | (19,468 – 46,794) | (2.7 - 6.5) |
| **Lifetime – full impacts** |  |  |  |  |  |  |
| NT 1c | 38,597 | 8.3 | 6,898 | 2.7 | 45,494 | 6.3 |
|  | (18,634 – 61,472) | (4.0 - 13.2) | (1,499 – 14,800) | (0.6 - 5.7) | (24,093 – 70,230) | (3.3 - 9.7) |
| NT 2c | 43,912 | 9.4 | 9,469 | 3.7 | 53,381 | 7.4 |
|  | (24,955 – 69,922) | (5.4 - 15.0) | (3,111 – 18,157) | (1.2 - 7.0) | (32,009 – 80,630) | (4.4 - 11.1) |
| WT c | 51,519 | 11.1 | 12,999 | 5.0 | 64,518 | 8.9 |
|  | (30,609 – 75,542) | (6.6 - 16.2) | (6,182 – 21,890) | (2.4 - 8.5) | (41,512 – 90,904) | (5.7 - 12.5) |

### Changes in incidence of stomach cancer by 2025 and 2030

Our model estimates that a gradual reduction in salt intake over six years to an average of 8g/day by 2025, could prevent about 2,900 (95% uncertainty interval [UI]: 1,109 to 5,199) new cases of stomach cancer in men and about 600 (95%UI: 133 to 1,316) in women, which translates to 6.2% and 2.8% relative reductions, respectively. In addition, over 11 years (by 2030) about 6,354 (95% UI: 2,790 to 11,030) and 1,346 (95% UI: 354 to 2,688) new cases of stomach cancer in men and women respectively could be prevented if the 7g/day national policy were achieved, that is, 7.3% and 3.3% relative reduction respectively. Furthermore, if the WHO target was attained by 2030, there could be 7,546 (95%UI: 3,969 to 11,916) and 1,732 (95%UI: 682 to 3,133) fewer incident cases of stomach cancer (8.6% and 4.3% reduction) for men and women, respectively. (Table 1).

### Changes in mortality from stomach cancer by 2025 and 2030

Over a period of six years (2019 – 2025), our model estimates that there could be 1,568 (95%UI: 526 to 2,959) and 383 (95%UI: 73 to 846) fewer deaths from stomach cancer men and women, respectively, that is, 3.0% and 1.5% relative reductions, if the mean population salt intake decreased linearly to 8g/day. Over a period of 11 years, that is, by 2030, achieving the 7g/day policy target could avert 4,231 (95%UI: 1,659 to 7,664) and 983 (95%UI: 239 to 2,024) stomach cancer deaths in men and women, respectively. Corresponding deaths avoided if the WHO target was achieved by 2030 were 5,065 (95%UI: 2,456 to 8,347) for men and 1,242 (95%UI: 464 to 2,302) for women. (Table 2).

**Table 2:** Estimated reduction in mortality from stomach cancer for adults in Vietnam between 2019 to 2030 and lifetime.

| Deaths | Men |  | Women |  | Both |  |
| --- | --- | --- | --- | --- | --- | --- |
|  | *Absolute reduction*<br>*n (95% UI)* | *Relative reduction*<br>*Est.% (95% UI)* | *Absolute reduction*<br>*n (95% UI)* | *Relative reduction*<br>*Est.% (95% UI)* | *Absolute reduction*<br>*n (95% UI)* | *Relative reduction*<br>*Est.% (95% UI)* |
| NT 1a – 2025 | 1,568 | 3.0 | 383 | 1.5 | 1,950 | 2.5 |
|  | (526 – 2,959) | (1.0 - 5.6) | (73 – 846) | (0.3 - 3.4) | (809 – 3,410) | (1.0 - 4.4) |
| NT 2a – 2030 | 4,231 | 4.7 | 983 | 2.3 | 5,214 | 3.9 |
|  | (1,659 – 7,664) | (1.8 - 8.5) | (239 – 2,024) | (0.6 - 4.8) | (2,428 – 8,855) | (1.8 - 6.7) |
| WT a – 2030 | 5,065 | 5.6 | 1,242 | 2.9 | 6,307 | 4.8 |
|  | (2,456 – 8,347) | (2.7 - 9.3) | (464 – 2,302) | (1.1 - 5.5) | (3,513 – 9,741) | (2.7 - 7.4) |
| **Lifetime with decay effects** |  |  |  |  |  |  |
| NT 1b – 2045 | 14,186 | 3.2 | 2,726 | 1.1 | 16,912 | 2.5 |
|  | (6,392 – 24,214) | (1.4 - 5.5) | (617 – 5,981) | (0.3 - 2.4) | (8,368 – 26,751) | (1.2 - 3.9) |
| NT 2b – 2050 | 20,009 | 4.5 | 4,231 | 1.7 | 24,240 | 3.5 |
|  | (9,391 – 32,940) | (2.1 - 7.4) | (1,228 – 8,359) | (0.5 - 3.4) | (12,737 – 37,916) | (1.8 - 5.5) |
| WT b – 2050 | 24,225 | 5.5 | 5,305 | 2.2 | 29,530 | 4.3 |
|  | (13,942 – 36,782) | (3.2 - 8.3) | (1,810 – 9,538) | (0.7 - 3.9) | (18,016 – 42,964) | (2.6 - 6.2) |
| **Lifetime – full impacts** |  |  |  |  |  |  |
| NT 1c | 34,533 | 7.8 | 6,263 | 2.5 | 40,796 | 5.9 |
|  | (16,765 – 55,035) | (3.8 - 12.4) | (1,376 – 13,462) | (0.6 - 5.5) | (21,455 – 63,032) | (3.1 - 9.1) |
| NT 2c | 39,234 | 8.9 | 8,585 | 3.5 | 47,819 | 6.9 |
|  | (22,190 – 62,618) | (5.0 - 14.1) | (2,775 – 16,555) | (1.1 - 6.7) | (28,749 – 72,460) | (4.2 - 10.5) |
| WT c | 46,059 | 10.4 | 11,787 | 4.8 | 57,846 | 8.4 |
|  | (27,422 – 67,613) | (6.2 - 15.3) | (5,597 – 19,862) | (2.3 - 8.1) | (37,253 – 81,791) | (5.4 - 11.9) |

### Estimated gains in healthy life years by 2025 and 2030

The above epidemiological changes in stomach cancer burden were projected to accrue extra 3,455 (95%UI: 1,058 to 6,662) HALYs by 2025, 13,816 (95%UI: 4,819 to 26,132) HALYs by 2030 (for the 7g/day target) and up to 16,213 (95%UI: 6,934 to 28,498) HALYs by 2030 (for the 5g/day target) in men. For women, corresponding benefits estimated were 919 (95%UI: 164 to 2,409), 3,515 (95%UI: 762 to 7,561) and 4,253 (95%UI: 1,364 to 8,265) HALYs (Table 3).

**Table 3:** Estimated HALYs gained between 2019 and 2030 and over the lifetime of adults in Vietnam.

|  | **Male** | **Female** | **Both** |
| --- | --- | --- | --- |
|  | **N (95% UI)** | **N (95% UI)** | **N (95% UI)** |
| NT1a – 2025 | 3,455<br>(1,058 – 6,662) | 919<br>(164 – 2,049) | 4,374<br>(1,729 – 7,736) |
| NT2a – 2030 | 13,816<br>(4,819 – 26,132) | 3,515<br>(762 – 7,561) | 17,332<br>(7,700 – 30,679) |
| WTa – 2030 | 16,213<br>(6,934 – 28,498) | 4,253<br>(1,364 – 8,265) | 20,466<br>(10,310 – 33,335) |
| **Lifetime with decay impact** |  |  |  |
| NT 1b – 2045 | 155,084<br>(71,178 – 255,081) | 32,106<br>(8,298 – 69,051) | 187,190<br>(95,622 – 291,201) |
| NT 2b – 2050 | 211,020<br>(102,955 – 339,893) | 47,886<br>(15,016 – 95,024) | 258,906<br>(141,652 – 399,052) |
| WT b – 2050 | 254,607<br>(149,340 – 377,825) | 60,904<br>(22,261 – 107,242) | 315,511<br>(199,811 – 451,842) |
| **Lifetime – full impact** |  |  |  |
| NT 1c | 301,295<br>(149,627 – 483,176) | 57,827<br>(12,607 – 122,470) | 359,122<br>(195,822 – 553,258) |
| NT 2c | 338,203<br>(189,358 – 533,870) | 78,053<br>(26,148 – 150,854) | 416,256<br>(254,095 – 627,671) |
| WT c | 398,610<br>(241,205 – 586,770) | 106,725<br>(50,002 – 179,881) | 505,336<br>(325,701 – 712,039) |

**Table 4:** Comparison of relative change in base case estimates from varying the TMREL for sodium in the univariate sensitivity analysis.

| Outcome | Scenarios | Base case | One-way sensitivity analysis |  |
| --- | --- | --- | --- | --- |
|  |  | TMREL 3g sodium<br>*Estimate (95% UI)* | TMREL 1g sodium<br>*Change from base* | TMREL 5g sodium<br>*Change from base* |
| **Incidence** | NT 1a-2025 | 3,499 (1,589 – 5,840) | +9.0% | -67.4% |
|  | NT 2a-2030 | 7,701 (3,880 – 12,684) | +14.3% | -71.9% |
|  | WT a-2030 | 9,278 (5,431 – 13,959) | +35.2% | -75.7% |
|  | NT 1b-2045 | 18,302 (9,103 – 28,999) | +8.1% | -68.3% |
|  | NT2b-2050 | 26,310 (13,892 – 41,175) | +15.4% | -73.0% |
|  | WT b-2050 | 32,046 (19,468 – 46,794) | +35.1% | -76.9% |
|  | NT1c-Lifetime | 45,494 (24,093 – 70,230) | +13.5% | -74.0% |
|  | NT2c-Lifetime | 53,381 (32,009 – 80,630) | +25.2% | -77.5% |
|  | WT c-Lifetime | 64,518 (41,512 – 90,904) | +62.6% | -79.9% |
| **Deaths** | NT 1a-2025 | 1,950 (809 – 3,410) | +7.6% | -65.0% |
|  | NT 2a-2030 | 5,214 (2,428 – 8 855) | +11.9% | -70.2% |
|  | WT a-2030 | 6,307 (3,513 – 9,741) | +27.5% | -74.4% |
|  | NT 1b-2045 | 16,912 (8,368 – 26,751) | +8.2% | -68.4% |
|  | NT2b-2050 | 24,240 (12,737 – 37,916) | +15.4% | -73.0% |
|  | WT b-2050 | 29,530 (18,016 – 42 964) | +35.3% | 77.0% |
|  | NT1c-Lifetime | 40,796 (21,455 – 63,032) | +13.3% | 74.0% |
|  | NT2c-Lifetime | 47,819 (28,749 – 72,460) | +25.1% | 77.4% |
|  | WT c-Lifetime | 57,846 (37,253 – 81,791) | +62.4% | 79.9% |
| **HALY** | NT 1a-2025 | 4,374 (1,729 – 7,736) | +7.1% | -66.6% |
|  | NT 2a-2030 | 17,332 (7,700 – 30,679) | +9.6% | -70.4% |
|  | WT a-2030 | 20,466 (10,310 – 33,335) | +21.1% | -73.6% |
|  | NT 1b-2045 | 187,190 (95,622 – 291,201) | +8.4% | -68.5% |
|  | NT2b-2050 | 258,906 (141,652 – 399,052) | +15.6% | -73.1% |
|  | WT b-2050 | 315,511 (199,811 – 451,842) | +36.7% | -77.1% |
|  | NT1c-Lifetime | 359,122 (195,822 – 553,258) | +13.2% | -73.6% |
|  | NT2c-Lifetime | 416,256 (254,095 – 627,671) | +24.5% | -76.9% |
|  | WT c-Lifetime | 505,336 (325,701 – 712,039) | +60.4% | -79.6% |

### Lifetime impacts and sensitivity analyses

In the scenarios with full impacts, that is, salt reduction policy targets achieved and sustained for the remaining lifecourse, our model projects that overall, about 45,494 (95%UI: 24,093 to 70,230), 53,381 (95%UI: 32,009 to 80,630) and 64,518 (95%UI: 41,512 to 90,904) new cases of stomach cancer could be averted at the 8g/day, 7g/day and 5g/day targets respectively. Corresponding stomach cancer deaths postponed were 40,796 (95%UI: 21,455 to 63,032), 47,819 (95%UI: 28,749 to 72,460) and 57,846 (95%UI: 37,253 to 81,791). In scenarios with a gradual phase out with nil policy effect beyond 2045 (for 8g/day target) and 2050 (for 7g/day and 5g/day targets), our model estimated 18,302 (95%UI: 9,103 to 28,999), 26,310 (95%UI: 13,892 to 41,175) and 32,046 (95%UI: 19,468 to 46,794), respectively for incident stomach cancers prevented and 16,912 (95%UI: 8,368 to 26,751), 24,240 (95%UI: 12,737 to 37,916) and 29,530 (95%UI: 18,016 to 42,964), respectively for deaths averted. With respect to healthy life years, the model estimates that an extra 359,000 (for the 8g/day target), 416,000 (for the 7g/day target) and over 505,000 (for the 5g/day target) stomach cancer-related HALYs could be gained. For the scenarios with a gradual phase out of policy effects, the corresponding estimates were 187,000 HALYs, 259,000 and over 315,000 HALYs gained (See Tables 1, 2 and 3)

In one-way sensitivity analysis, compared to a TMREL of 3g of sodium (base case), using a TMREL of 1g of sodium resulted in larger health gains in the order of 7 - 9% for the 8g/day target by 2025. In addition, by 2030, outcomes increased by 9 – 14% (for the 7g/day target) and 21 – 35% (for the WHO target) compared to base case. For the lifetime scenarios, an optimal intake of 1g of sodium increased health gains observed by 13 – 25% relative to base case for the national targets and up to 62% for the WHO target. On the other hand, considering a higher TMREL of 5g of sodium considerably reduced the health benefits by 65 – 79% for all targets and across the short-term and lifetime horizons (See Figure 2).

**Figure 2:**
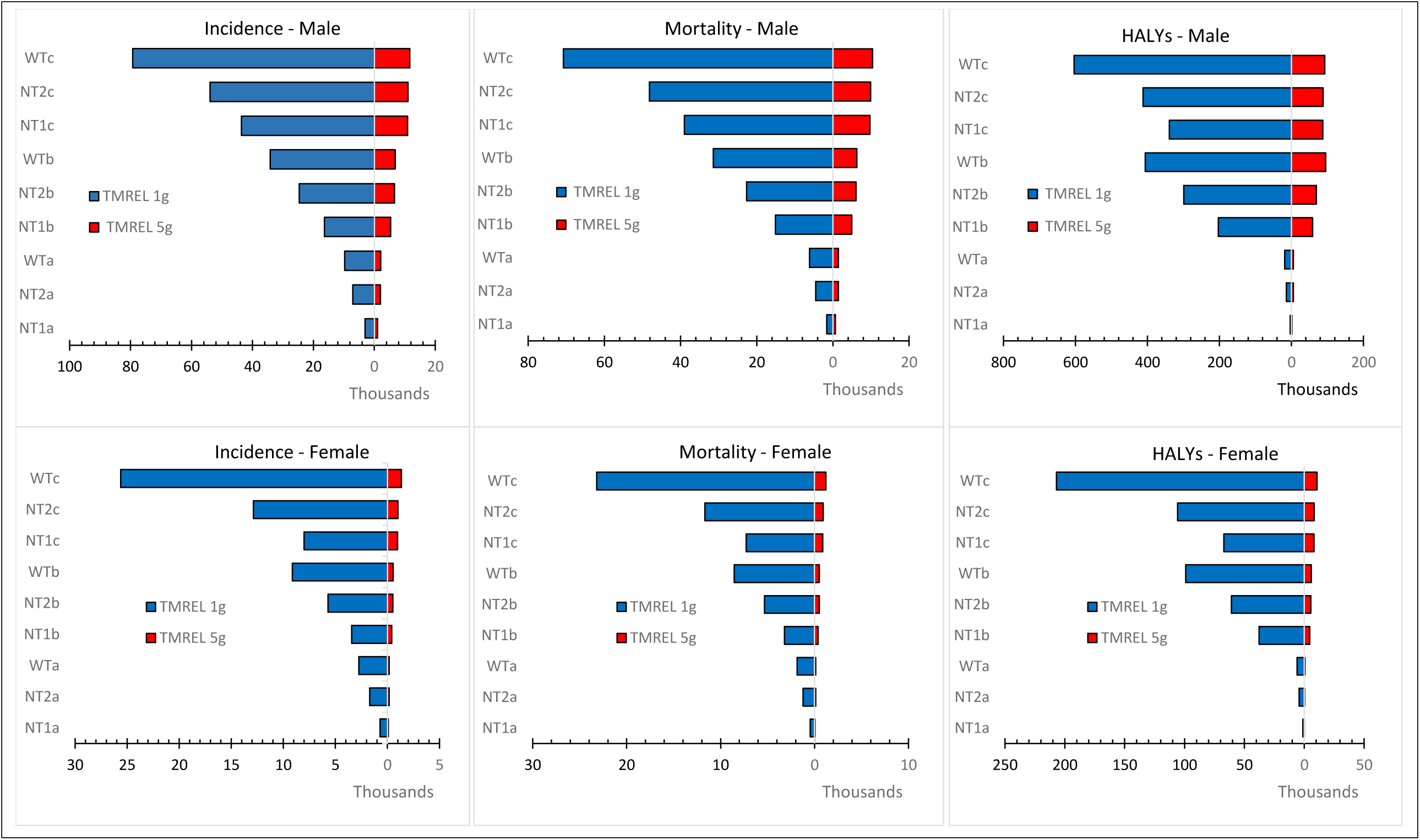
Tornado plot of one-way sensitivity analysis showing the impacts different TMRELs on incidence, mortality and HALYs over varied time horions. TMREL, Theoretical minimum risk exposure level (optimal level of consumption at which disease risk is lowest); *NT1a*, National target of 8g/day salt intake achieved by 2025; *NT2a*, National target of 7g/day salt intake achieved by 2030; *WTa*, WHO target of 5g/day achieved by 2030; *NT1b*, National target of 8g/day achieved by 2025 and effect gradually phased out to zero impact beyond 2045; *NT2b*, National target of 7g/day achieved by 2030 and effect gradually phased out to zero impact beyond 2050; *WTb*, WHO target of 5g/day achieved by 2030 and effect gradually phased out to zero impact beyond 2050; *NT1c*, National target of 8g/day achieved and effect sustained over lifetime; *NT2c*, National target of 7g/day achieved and effect sustained over lifetime; *WTc*, WHO target achieved and effect sustained over lifetime.

## Discussion

In this study, we estimate the potential impact of achieving the 2018 Healthy Vietnam Program salt reduction targets on the future burden of stomach cancer and population health. Compared to current levels of salt consumption, 3,500 incident cases of and 1,950 deaths from stomach cancer could be prevented if the 8g/day target was achieved over six years, that is, by 2025. Achieving the 7g/day target over 11 years (by 2030) could avert 7,700 new cases of and 5,200 deaths from stomach cancer. These changes in stomach cancer burden translated to 4,374 and 17,332 HALYs gained by 2025 and 2030, respectively. Larger impacts in avoidable disease burden could be obtained with further reductions in salt intake down to the WHO target of 5g/day, for which we estimate over 6,300 stomach cancer deaths prevented or postponed and 20,466 HALYs gained by 2030. Similarly, if these reductions in salt consumption to the national and WHO targets were sustained over the remaining lifetime, even bigger health gains could be attained. Our results were sensitive to varied optimal levels of salt intake. As expected, larger impacts were obtained with an optimal intake level 1g of sodium as opposed to considerable shrinking when a higher optimal intake level of 5g of sodium was used.

Very few studies (mostly from high-income countries) have quantified the long-term impacts of excess salt intake on the burden of stomach cancer. Kypridemos et al. modelled various scenarios in relation to the well-known United Kingdom 2003 – 2011 salt reduction program. This program included public awareness campaigns, food labelling and voluntary reformulation of processed foods by food industries. In a scenario where the decline in salt intake observed in the program continued to 2015 (that is, 8.9g/day in 2003 to 7.1g/day in 2015), an estimated 5,000 incident cases of stomach cancer and 2,000 fewer deaths from the disease were prevented over that period (22). Further salt reductions to ∼6 g/day by 2030 were estimated to result in an additional 1,200 new stomach cancer cases prevented and 700 deaths postponed. Another recent modelling study in the United States has shown that compared to status quo, a low-sodium DASH diet could reduce the lifetime risk of stomach cancer by 24.8% in men and 21.2% in women. In addition, this prevented 27 and 14 stomach cancer cases per 10,000 people respectively for men and women and postponed 24 stomach cancer deaths in men and 13 stomach cancer deaths in women per 10,000 persons (23). Despite differences in modelling approaches, with Kypridemos et al. using dynamic microsimulation and the latter study using Markov models, the results from these studies are akin to our findings - demonstrating the substantial avoidable stomach cancer burden from population-wide salt reduction.

Our results show that benefits were significantly greater in men than women. This could be explained by several factors. First, men had much higher baseline salt intake and hence needed a greater absolute salt reduction to meet the program targets modelled. Secondly, men have higher incidence (nearly twice for some age-groups) and slightly higher fatality rates (in older ages) for stomach cancer compared to women. The high hazard rates in men are consistent with the recent 2020 GLOBOCAN report (3). It is in part explained by the clustering of other major risk factors for gastric cancer such as smoking and heavy alcohol use in men. Put together, it is highly likely that men would benefit more from primary prevention interventions that modify population exposures by shifting the distributions of major risk factors like salt intake to healthy ranges.

### Public Health and policy implications

Over two-thirds of adult Vietnamese reported that they consume *just the right amount* of salt. However, estimates from urinary sodium excretion indicate actual intake is nearly twice that recommended by the WHO (10). As most (70%) of the salt consumed in Vietnam is added during cooking or at the table and from fish sauces (10, 24), strategies to achieve the Healthy Vietnam Program salt reduction targets need to focus on consumers. Do and colleagues evaluated a suite of behavioural change interventions including mass media, school interventions and community programs conducted for a year, and found significant reductions in sodium intake in a province in Vietnam (25). Such interventions could be scaled nation-wide across all 63 provinces to achieve larger returns. Recently, the government of Vietnam through the Department of Preventive Medicine of the Ministry of Health in collaboration with the WHO country office and support from Vital Strategies organization have commenced a nation-wide media campaign to educate the public on the ills of excess salt intake and the need to reduce its consumption (24). The longterm sustainability of these programs is unknown. However, recent modelling assessing the lifetime impacts of media campaigns and school-based programs for salt reduction found that despite the modest effects, these interventions could deliver large population health gains and were cost-saving if effects are sustained (26).

For Vietnam and other LMICs where most of the salt is added during cooking, another viable intervention for consideration is replacing regular salt with low-sodium potassium-rich salt-substitutes. Evidence from large pragmatic community trials have shown that they are very effective in reducing blood pressure and vascular events (27, 28), as well as being highly cost-effective (26). Nonetheless, given the expansion of multinational food industries in many LMICs, partly fuelling the nutrition transition with growing consumption of ultra-processed foods rich in salt, sugar, and fats, these governments will need to consider multi-pronged strategies. Among others, the inclusion of upstream interventions like industry food reformulation, food labelling using consumer-friendly methods that are easy to interpret like traffic light warnings, would be needed (29). These measures must be accompanied by regular surveillance, monitoring and evaluation to assess progress and identify areas needing attention to ensure sustained and equitable impacts.

Recent analysis of the burden of gastric cancer showed that the age-standardized incidence and death rates have significantly declined since 1990. However, the rate of decline appears to have slowed down and plateaued in recent years, especially in East Asia and Pacific region (2, 30). It is suggested that this decline may partially be explained by declining trends in the global prevalence of smoking (another major risk factor for stomach cancer) (30). We posit that the recent plateau (and reversal for some countries in Asia) in the trend might in part be driven by increasing salt consumption that has accompanied the nutrition transition in Asia (31, 32), which has the highest salt consumption levels globally (20). Moreover, the GBD study showed that while about 38.2% of global age-standardized stomach cancer-related DALYs were attributable to diets high in salt (smoking was second contributor at 24.5%), a much higher fraction (61.3%) was observed in East Asia, approximately twice that of other regions (2). This demonstrates the substantial contribution that high salt diets in the region make to the stomach cancer-related morbidity and mortality.

During the last two decades, the government of Vietnam has made commitments towards cancer control, for example, setting up the National Cancer Control Program as part of the national strategy for the prevention and control of NCDs 2015-2025 (33). However, a recent review highlighted a number of shortcomings including limited availability and accessibility to diagnostic and treatment services across some provinces; inadequate funding (2.5-3.5% of total health expenditure) for NCD programs despite NCDs accounting for 70% of all deaths; low community awareness with nearly four-fifths of patients diagnosed at advanced stages of cancer; and limitations in data infrastructure (34). The high lethality of stomach cancer, limited capacity and associated healthcare costs underscore the need for primary prevention to take precedence in efforts to combat this condition. Simple effective and cost-effective strategies like population dietary salt reduction should therefore be high on the policy agenda for measures to mitigate the burden of stomach cancer in Vietnam, Asia and globally.

### Strengths and limitations

Our study has some limitations. First, our modelling does not consider the future trends in salt consumption. Given the ongoing nutrition transition, salt intake may increase over time. It is also possible that consumption might decline in response to the recent government initiatives. Predicting the balance of this is not immediately straightforward, however, our analysis reveals that with the current high salt consumption, even very modest reductions in salt intake are likely to deliver sizeable health gains. Second, we do not explicitly model the lag time from exposure to incidence of stomach cancer. This implies our estimates maybe on the high end, however, in our lifetime scenarios, we model intervention decays down to zero impact beyond 20 years, which mitigates any overestimation. Third, we model a closed cohort of adult Vietnamese. Thus, our lifetime health outcomes do not capture the benefits of salt reduction that accrue from younger cohorts as they age out, rendering our results conservative.

Despite the above, our study has some strengths. To our knowledge this study is the first from Asia to estimate the burden of stomach cancer that could be avoided from dietary salt reduction. Our findings contribute to bridging the knowledge gap in the region and provides important evidence needed by policy makers as they prioritise strategies to tackle cancer and NCDs. Furthermore, by quantifying the differential impact of varied TMRELs, we demonstrate that irrespective of the optimal salt intake level considered, substantial gains in population health could be obtained. Finally, we model multiple time horizons including lifecourse, which goes beyond the current national targets and thereby provides comprehensive evidence on the long-term impacts of policy actions or inactions. Finally, by using local survey data on salt consumption, we allay any issues with transferability of evidence from elsewhere, given that most of the evidence on future impacts of salt intake on stomach cancer is from high-income countries.

## Conclusion

Our study suggests that achieving the Healthy Vietnam program salt reduction targets could avert a substantial proportion of the future stomach cancer burden in Vietnam by 2025 – 2030 and gain thousands of healthy life years. Larger health benefits could be obtained if the WHO salt reduction target was met, and targets sustained over the lifespan. Given the high lethality of stomach cancer and limited diagnostic and therapeutic resources in Vietnam, investing in primary prevention strategies such as salt reduction should be high on the policy agenda to optimise population health and alleviate future pressure on the health system.

## Data Availability

All data produced in the present work are contained in the manuscript. Data from from the Global Burden of Disease Study used in this project are publicly available at: http://ghdx.healthdata.org/gbd-results-tool

