## Supplementary file for "Avoidable burden of stomach cancer and potential gains in healthy life years from gradual reductions in salt consumption in Vietnam, 2019 – 2030: a modelling study"

**Input data**

**Table 1: Salt intake levels and estimated annual reductions in mean salt intake towards the 2025 and 2030 targets**

|  | Current levels ^[1]^ for base year 2019 | National Target 1 to 2025 | National Target 2 to 2030 | WHO Target to 2030 |
| --- | --- | --- | --- | --- |
| Men | Mean (SD) | Mean | Mean | Mean |
| 18 – 30 years | 10.3 (2.4) | 0.383 | 0.300 | 0.482 |
| 31 – 49 years | 10.7 (2.9) | 0.450 | 0.336 | 0.518 |
| 50 – 69 years | 10.6 (2.7) | 0.433 | 0.327 | 0.509 |
| Women |  |  |  |  |
| 18 – 30 years | 8.2 (2.3) | 0.033 | 0.109 | 0.291 |
| 31 – 49 years | 8.6 (2.2) | 0.100 | 0.145 | 0.327 |
| 50 – 69 years | 7.9 (2.4) | 0.000 | 0.082 | 0.264 |

**Table 2: Relative risk, incidence, and case fatality rates of stomach cancer in Vietnam in 2019**

| **Parameter** | **Category** | **Men** | **Women** | **Source** |
| --- | --- | --- | --- | --- |
| Relative risks* | All ages | 1.33 (1.04 – 1.43) | 1.33 (1.04 – 1.43) | GBD 2019 ^[2]^ |
| Incidence rates | 25 – 29 years | 0.0000 | 0.0000 | Outputs from DISMOD-II based on GBD 2019 data for Vietnam |
|  | 30 – 34 years | 0.0000 | 0.0000 |  |
|  | 35 – 39 years | 0.0000 | 0.0000 |  |
|  | 40 – 44 years | 0.0001 | 0.0000 |  |
|  | 45 – 49 years | 0.0001 | 0.0001 |  |
|  | 50 – 54 years | 0.0002 | 0.0001 |  |
|  | 55 – 59 years | 0.0004 | 0.0001 |  |
|  | 60 – 64 years | 0.0006 | 0.0002 |  |
|  | 65 – 69 years | 0.0008 | 0.0003 |  |
|  | 70 – 74 years | 0.0009 | 0.0003 |  |
|  | 75 – 79 years | 0.0011 | 0.0004 |  |
|  | 80 – 84 years | 0.0013 | 0.0005 |  |
|  | 85 – 89 years | 0.0018 | 0.0006 |  |
|  | 90 – 94 years | 0.0018 | 0.0007 |  |
|  | 95+ years | 0.0017 | 0.0007 |  |
| Case fatality rates | 25 – 29 years | 0.0355 | 0.0627 | Outputs from DISMOD-II based on GBD 2019 data for Vietnam |
|  | 30 – 34 years | 0.1972 | 0.2643 |  |
|  | 35 – 39 years | 0.3193 | 0.3667 |  |
|  | 40 – 44 years | 0.3549 | 0.3891 |  |
|  | 45 – 49 years | 0.3596 | 0.4007 |  |
|  | 50 – 54 years | 0.3876 | 0.4185 |  |
|  | 55 – 59 years | 0.4433 | 0.4554 |  |
|  | 60 – 64 years | 0.5258 | 0.5342 |  |
|  | 65 – 69 years | 0.6095 | 0.6098 |  |
|  | 70 – 74 years | 0.7388 | 0.7103 |  |
|  | 75 – 79 years | 0.8992 | 0.8526 |  |
|  | 80 – 84 years | 1.0772 | 1.0404 |  |
|  | 85 – 89 years | 1.3695 | 1.3240 |  |
|  | 90 – 94 years | 1.8321 | 1.7368 |  |
|  | 95+ years | 1.9976 | 1.9521 |  |

*****Relative risks of stomach cancer per 3g/day increment in sodium.

**Table 3: Prevalence and disability rates of stomach cancer in Vietnam in 2019**

| **Parameter** | **Category** | **Men** | **Women** | **Source** |
| --- | --- | --- | --- | --- |
| Prevalence rates | 25 – 29 years | 0.0000 | 0.0000 | Outputs from DISMOD-II based on GBD 2019 data for Vietnam |
|  | 30 – 34 years | 0.0001 | 0.0000 |  |
|  | 35 – 39 years | 0.0001 | 0.0001 |  |
|  | 40 – 44 years | 0.0002 | 0.0001 |  |
|  | 45 – 49 years | 0.0003 | 0.0001 |  |
|  | 50 – 54 years | 0.0005 | 0.0002 |  |
|  | 55 – 59 years | 0.0008 | 0.0002 |  |
|  | 60 – 64 years | 0.0010 | 0.0003 |  |
|  | 65 – 69 years | 0.0012 | 0.0004 |  |
|  | 70 – 74 years | 0.0013 | 0.0005 |  |
|  | 75 – 79 years | 0.0012 | 0.0005 |  |
|  | 80 – 84 years | 0.0012 | 0.0005 |  |
|  | 85 – 89 years | 0.0013 | 0.0005 |  |
|  | 90 – 94 years | 0.0010 | 0.0004 |  |
|  | 95+ years | 0.0009 | 0.0004 |  |
| Stomach cancer disability rates | 25 – 29 years | 0.12740 | 0.14107 | Calculated from GBD 2019 data.  **[ DW =** Stomach Ca YLD #s**/** # Prev Stomach Ca **]**  **[ AdjDW = DW**/ (1- (All-cause YLD #s -Stomach Ca YLD #s) **/** Pop #) **]**  #= numbers, DW=disability weight, Adj= adjusted. |
|  | 30 – 34 years | 0.13670 | 0.15112 |  |
|  | 35 – 39 years | 0.15057 | 0.16307 |  |
|  | 40 – 44 years | 0.15646 | 0.17092 |  |
|  | 45 – 49 years | 0.16345 | 0.17211 |  |
|  | 50 – 54 years | 0.17200 | 0.17661 |  |
|  | 55 – 59 years | 0.17464 | 0.18311 |  |
|  | 60 – 64 years | 0.18369 | 0.19864 |  |
|  | 65 – 69 years | 0.20117 | 0.22111 |  |
|  | 70 – 74 years | 0.23040 | 0.23348 |  |
|  | 75 – 79 years | 0.27284 | 0.26795 |  |
|  | 80 – 84 years | 0.30375 | 0.31814 |  |
|  | 85 – 89 years | 0.35925 | 0.38260 |  |
|  | 90 – 94 years | 0.42945 | 0.46590 |  |
|  | 95+ years | 0.51149 | 0.60148 |  |
| All-cause disability rates | 25 – 29 years | 0.0619 | 0.0880 | Calculated from GBD 2019 age- & sex-specific data for Vietnam  **[ pYLD =** All-cause YLD numbers / Population numbers **]** |
|  | 30 – 34 years | 0.0727 | 0.0985 |  |
|  | 35 – 39 years | 0.0803 | 0.1084 |  |
|  | 40 – 44 years | 0.0873 | 0.1189 |  |
|  | 45 – 49 years | 0.0975 | 0.1282 |  |
|  | 50 – 54 years | 0.1102 | 0.1390 |  |
|  | 55 – 59 years | 0.1264 | 0.1525 |  |
|  | 60 – 64 years | 0.1458 | 0.1742 |  |
|  | 65 – 69 years | 0.1716 | 0.2034 |  |
|  | 70 – 74 years | 0.2053 | 0.2374 |  |
|  | 75 – 79 years | 0.2413 | 0.2707 |  |
|  | 80 – 84 years | 0.2729 | 0.3090 |  |
|  | 85 – 89 years | 0.3005 | 0.3410 |  |
|  | 90 – 94 years | 0.3185 | 0.3680 |  |
|  | 95+ years | 0.3331 | 0.4008 |  |
